# Classifying the risk for myasthenic crisis using data-driven explainable machine learning with informative feature design and variance control – a pilot study

**DOI:** 10.1101/2023.08.19.23294175

**Authors:** Sivan Bershan, Andreas Meisel, Philipp Mergenthaler

## Abstract

**Importance:** Myasthenic crisis (MC) is a critical progression of Myasthenia gravis (MG), requiring intensive care treatment and invasive therapies. Classifying patients at high-risk for MC facilitates treatment decisions and helps prevent disease progression.

**Objective:** To test whether machine learning models trained with real-world routine clinical data can aid precisely identifying MG patients at risk for MC.

**Design:** This is a pseudo-prospective cohort study of MG patients presenting since January 2010.

**Setting:** Single center.

**Participants:** A cohort of 51 MG patients was used for model training based on a defined set of real-world clinical data. The cohort was created from a convenience sample of 13 MC patients matched based on sex, five-year age band, antibody status, thymus pathology with MG patients who had not suffered an MC. Data analyses and model refinements were performed from June 2022 to May 2023.

**Exposure:** Classification of MG patients to high or low risk for MC using Lasso regression or random forest machine learning models.

**Main Outcomes and Measures:** The accuracy of the risk classification was assessed by patient.

**Results:** This study included 51 MG patients (13 MC, 38 non-MC; median age MC group 70.5, non-MC group 65.5). The mean cross-validated AUC classifying MG patients as high or low risk for MC based on simple or compound features derived from real-world routine clinical data showed a predictive accuracy of 68.8% for the regularized Lasso regression and of 76.5% for the random forest model. Feature importance scores suggest that multimorbidity may play a role in risk classification. Different thresholds were applied to tune model performance to optimal parameters. Studying result stability across 100 runs further indicated that the random forest model was better suited to cope with feature variance. Studying feature importance across 5100 model runs identified explainable features to distinguish MG patients at high or low risk for MC.

**Conclusions and Relevance:** In this study, feasibility of classifying risk for MC based on real-world routine clinical data using machine learning was shown. The models showed accurate and consistent performance indicating the utility of personalized risk assessment in MG patients using machine learning models.

**Key Points:** *Question:* Can machine learning models be used to classify Myasthenia gravis patients into groups at high or low risk for myasthenic crisis with high precision based on explainable data-driven features derived from real-world clinical data?

*Findings:* In this pseudo-prospective study of 51 Myasthenia gravis patients, the risk of myasthenic crisis using real-world clinical data was accurately classified employing two machine learning models with explainable features.

*Meaning:* These findings suggest that it is possible to classify the risk for myasthenic crisis in patients based on real-world clinical data with high precision.

## Introduction

Myasthenia gravis (MG) is a rare chronic autoimmune disease causing fatigable muscle weakness due to auto-antibody-mediated decrease in neuromuscular transmission with a prevalence of 40-180 per 1 million people^1,2^. Myasthenic crisis (MC) defines critical exacerbation of MG which can be life threatening due to respiratory insufficiency and requires intensive care treatment, mechanical ventilation as well as invasive therapeutic procedures such as plasmapheresis. Up to 15-20% of MG patients develop MC over the course of their lifetime^3,4^. Although there has been a significant decrease in mortality of MC over the last decades, current figures for mortality are variable, but still reported as up to 5-12%^4–6^. Even though the presence of thymoma, MuSK autoantibodies^7^, stress, infections or inappropriate treatment are known risk factors for MC, among others, it is still impossible to anticipate MC or predict which patients develop MC in a cohort of patients at risk. The ability to predict which patients have a high risk of MC will help make confident and personalized treatment decisions and thereby help utilize resources more effectively.

In predictive modeling the objective is to accurately project the chances that a specific event will or will not happen, thereby optimizing for prediction accuracy and not for the understanding of root causes. While byproducts such as the feature importance can give insight into why an event occurs, the primary interest lies in predicting if it will occur^8^. In principle, two classes of models are suited for prediction: regression, generally used for predicting a continuous numeric outcome, and classification for categorical outcomes.

Here, we investigated whether it is possible to reliably classify Myasthenia gravis patients into groups at low- or high risk of MC based entirely on routine medical data in a proof-of-concept pseudo-prospective pilot study. Ultimately, our goal is to support making treatment decisions in a clinical context. Thus, we used real-world routine medical data such as common laboratory values and other case-associated data to classify patients from a pilot cohort of MG patients into MC risk groups. In an explainable data-driven approach, we investigated how to best classify patients into risk groups using either a regularized linear machine learning model to account for highly correlated features or a random forest classifier to minimize noise.

## Methods

### Protocol approval and patient consent

This study was approved by the local ethics committee (no. EA4/068/22). Informed consent was not required for this retrospective analysis.

### Study design and participants

This study is a pilot study to demonstrate feasibility of MC prediction based on real-world routine clinical data. In this study our dependent variable allowed for the two categories: “myasthenic crisis” or “no myasthenic crisis”. Thus, we treated this as a classification model.

We chose a 2-step approach in a pseudo-prospective manner (i.e., occurrence of MC was unknown to the machine learning models). First, we used univariate logistic regression to assess feature importance, and then we compared regularized regression with random forest classification to classify risk into low or high risk for MC. Details of model generation, performance testing and validation are given in the statistical analysis section below. In order to perform pseudo-prospective predictive analysis, we designed a cohort of MG patients from retrospective medical data of patients all treated since January 2010 until recent at the Integrated Myasthenia gravis Center of the Dept. of Neurology at Charité – Universitätsmedizin Berlin, a large academic tertiary care center, certified for applying standardized clinical pathways and patient management by the German Myasthenia Gravis Society. 53 patients with MC admitted to the neurological intensive care unit were screened, of which 13 were included in this pilot study (Table 1). To establish the final cohort for analysis, we initially matched each MC patient with up to four MG patients without MC based on (in order of priority) sex, five-year age band, antibody status, thymus pathology. After data cleanup (below), we were able to match 38 control patients.

**Table 1:** Clinical and demographic characteristics of patients.

| **Table 1:** Clinical and demographic characteristics of patients |  |  |  |
| --- | --- | --- | --- |
|  |  | **Myasthenic crisis** | **Control** |
| **Number of Patients** |  | 13 | 38 |
| **Age at time of sampling, median (range)** |  | 70.5 (39 – 89) | 65.5 (40 – 88) |
| **Disease duration (years), median (range)** |  | 13.5 (7-43) | 9.5 (4 – 43) |
| **Onset, n (%)** | Late Onset MG | 8 (61%) | 20 (52%) |
|  | Early Onset MG | 5 (38%) | 18 (47%) |
| **Sex, n (%)** | Female | 6 (46%) | 17 (45%) |
|  | Male | 7 (54%) | 21 (55%) |
| **Antibody status, n (%)** | AChR | 11* (85) | 27 (71) |
|  | AB-negative | 2 (15) | 11 (19) |
| **Thymectomy, n (%)** | No | 4 (31) | 11 (29) |
|  | Yes | 9 (69) | 27 (71) |
| **Thymus pathology, n (%)** | Thymoma | 5 (56) | 12 (44.5) |
|  | Hyperplasia | 1 (11) | 3 (8) |
|  | Unremarkable | 3 (33) | 12 (44.5) |
| **Patients with number of MC, n** | 0 MC | 0 | 38 |
|  | 1 MC | 9 | 0 |
|  | 2 MC | 2 | 0 |
|  | 3 MC | 2 | 0 |
| *\*One MC patient was positive for AChR and MuSK. As there were no appropriate AChR/MuSK-double positive controls she was considered in the AChR+ group.* |  |  |  |

MC was defined as exacerbation of myasthenic symptoms with bulbar or general weakness requiring mechanical ventilation. Diagnosis of MG was established based on antibody findings, repetitive nerve stimulation or clinical assessment. Current data analyses and model refinements were performed from June 2022 to May 2023.

### Data curation and preprocessing

All data were obtained from the patients’ electronic health records or from Charité’s Health Data Platform (HDP) which hosts up-to-date retrospective snapshots of the entire hospital management system, including laboratory values. To eliminate hindsight bias, we removed data that were generated within 6 months after the MC.

Input data were routine clinical data such as bloodwork, data relating to current hospital admission (e.g., length of stay, path through hospital, etc.), as well as treatment details (e.g., medication, procedures, etc.). A full list of considered features is shown in Supplementary Table 1. The data were cleaned from impossible values (e.g., negative laboratory values), normalized and standardized. The matching criteria were excluded as classifiers, since by design they were similar in test and control groups. Two patients with MC had to be excluded from the analysis because they had no remaining data after eliminating the data generated in the half year period after the crisis.

### Statistical and Machine Learning Analysis

#### Modeling Approach

Analyses were performed in R version 3.6.1 (R Project for Statistical Computing). For a full list of libraries used see Supplementary Table 2.

We used several feature categories for analyses: bloodwork, hospitalization, and treatment details (see Data Curation above for details). In regression models it is typical to use one row by patient and to depict trend in the feature design. An example is the number of encounter days by patient in general and the corresponding trend feature would be the average encounter days by patient per six months. If applicable, we added minimum, maximum, median, and standard deviation for each value as simple features. For features that did not change over time (e.g., age of onset), this was omitted. For the regularized regression, we also allowed pairwise interactions. In the random forest model, we allowed only simple features and no interactions, because the tree structure itself allows for nonlinear relationships. For the full list of features used see the labels in Fig. 3.

We set a minimum completeness level of 80% per feature, meaning that at least 41 patients had to have a value for a particular feature. Out of originally more than 2000 possible features, 696 feature candidates reached the completeness threshold to be considered in the models. The missing values for the features used were computed with the mice package^9^ using predictive mean matching for numeric features. This method predicts the value to be imputed based on all other values except the dependent variable. Then it draws a small set of candidate donors closest to the predicted value and draws one of these randomly^10^. Factor data follows the same process with the exception that the prediction is performed with a polyregression.

In a first step, we determined feature importance in a logistic regression model. Features with a p-value of ≤ 0.05 were then used in a regularized Least Absolute Shrinkage and Selection Operator (Lasso) regression^11^ to account for many features being highly correlated among the top 50 from the first step. The Lasso regression algorithm identified 8 - 11 features per run that were most predictive. The parameter λ controls the strength of the shrinkage, where an increase in λ results in an increase in shrinkage and an increase in variance. Due to the significant reduction in features, variance is introduced through the model. We thus also calculated a random forest model to gauge if variance was controlled well.

#### Performance Metrics and Validation

Our primary model performance metric in both second phase models was the mean of the cross-validated area under the receiver-operator curve (AUC) over 100 runs of training. AUC is a classification threshold independent metric, contrary to comparison metrics such as sensitivity and specificity which are highly dependent on what threshold is chosen to distinguish the groups.

Accounting for the small data set, we performed leave-one-out cross-validation^12^. Standard metrics such as sensitivity, specificity, and precision were used to evaluate model performance. We also ran 100 cycles of training to account for two sources of randomness – imputation and the L1-regularization. L1-regularization penalizes the sum of absolute values and is sparse, meaning it sets all variables but the top ones to zero and doesn’t use them. Finally, we scrambled the target variable and verified that the results had a significantly lower AUC.

### Data and code availability

Feature categories and lists are published as supplement to this manuscript (Supplementary Table 1). Ethical approval currently does not permit sharing of raw data. The analysis code will be made available upon reasonable request.

## Results

### Demographics and clinical characteristics

The cohort consisted of 51 Myasthenia gravis patients with 13 patients who suffered from at least one MC (9 patients had one MC, 4 had two or more MC) and 38 controls (Table 1). The median age in the MC group was 70.5, whereas the non-MC group showed a median age of 65.5. Overall, 38 patients were AChR antibody positive and the remaining 13 were antibody negative. One patient tested positive for both AChR as well as MuSK, who we matched against AChR single-positive patients due to a lack of other controls.

### Model Performance

The Lasso regression model allowed the distinction between the MC and non-MC groups with a mean [standard deviation, sd] AUC of 68.8% [8.1%] (Figure 1A, Table 2), and random forest with a mean AUC of 76.7% [4%] (Figure 1B, Table 2).

**Fig. 1:**
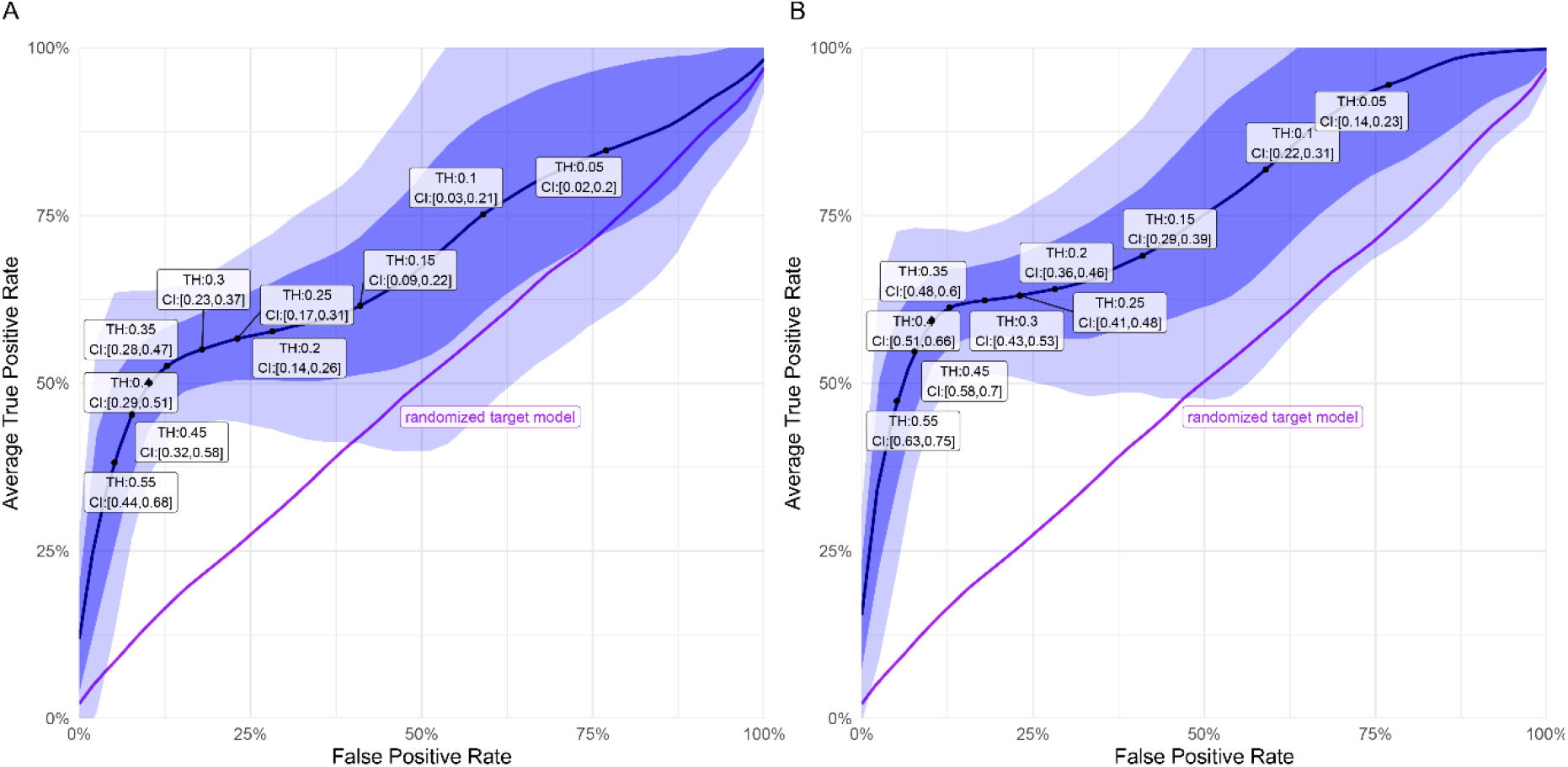
Prediction results independent of threshold (**A)** for the Lasso regression, and (**B**) for the random forest prediction. The random forest prediction performs better in terms of AUC. The black line is the area under the curve for the prediction, the shaded dark blue area represents one standard deviation confidence intervals (CI) and the light blue 2 standard deviations CI. The labels show the thresholds and the respective CI. The purple line represents the prediction with the randomized target variable.

**Table 2:**
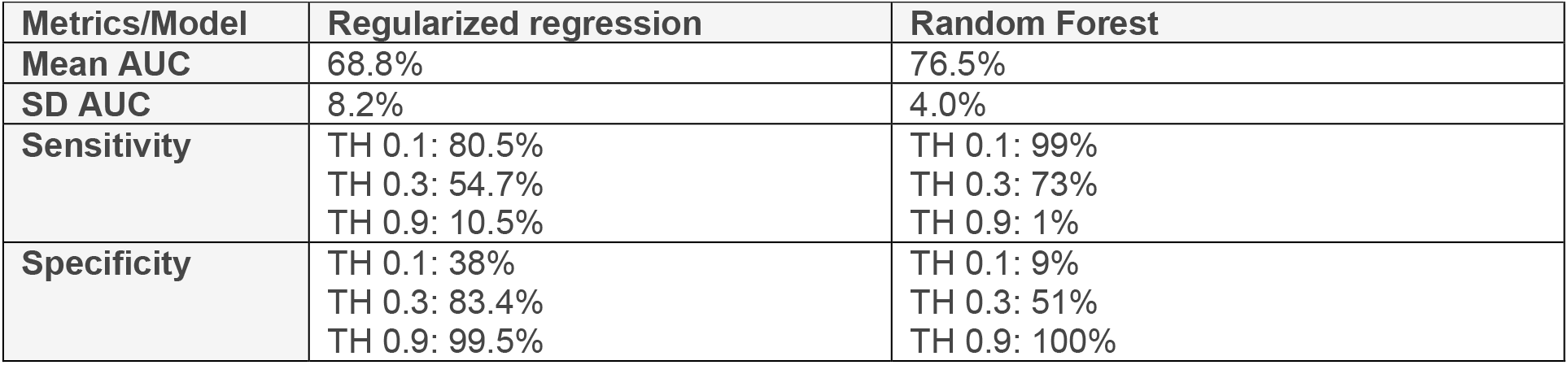
Result summary at different thresholds (TH)

| Table 2: Result summary at different thresholds (TH) |  |  |
| --- | --- | --- |
| Metrics/Model | Regularized regression | Random Forest |
| Mean AUC | 68.8% | 76.5% |
| SD AUC | 8.2% | 4.0% |
| Sensitivity | TH 0.1: 80.5%<br>TH 0.3: 54.7%<br>TH 0.9: 10.5% | TH 0.1: 99%<br>TH 0.3: 73%<br>TH 0.9: 1% |
| Specificity | TH 0.1: 38%<br>TH 0.3: 83.4%<br>TH 0.9: 99.5% | TH 0.1: 9%<br>TH 0.3: 51%<br>TH 0.9: 100% |

Three configuration examples of the Lasso regression model (Table 2) show that the best accuracy (i.e., classifying most patients correctly) is not what the model should be optimized for. It is our major aim to correctly identify patients at risk for MC. It is therefore critical to reduce false negatives, because false negatives mean that patients with high risk of suffering an MC would be classified as “low risk” and may be overlooked. We thus shifted the threshold splitting the two categories to reduce false negatives as much as possible. For this, we looked at the average predicted score by patient.

The confusion matrices for the Lasso regression prediction (Fig. 2A) show that at a threshold of 0.1, 35 out of 51 patients would be considered high risk for MC. Ten of these were correctly classified. The high number of 25 false positives was accompanied by a low number of false negatives (3 patients). Accordingly, at a threshold of 0.3, 13 patients were categorized as high risk, 7 of these true positives. 38 patients were considered low risk, whereas 6 patients were false negatives. In this setting, the number of false negatives doubled compared to the lowest threshold. To gauge the result range, at a threshold of 0.8, 50 patients were considered low risk, 1 patient was considered high risk, and 12 MC patients were false negatives. In the random forest model (Fig. 2B) and at a threshold of 0.1, 48 patients (94%) were considered high risk. 13 of these were correctly classified, whereas 0 patients were false negatives. At a threshold of 0.3, 29 patients (57%) were considered high risk. Of these, 9 were correctly classified and 4 patients were false negatives. At a threshold of 0.8, 0 patients (0%) were considered high risk. The goal is to maintain reasonable group sizes allowing the allocation of significantly more resources to those in high risk. False negatives should be avoided, even if this means that the precision of the model is lowered.

**Fig. 2:**
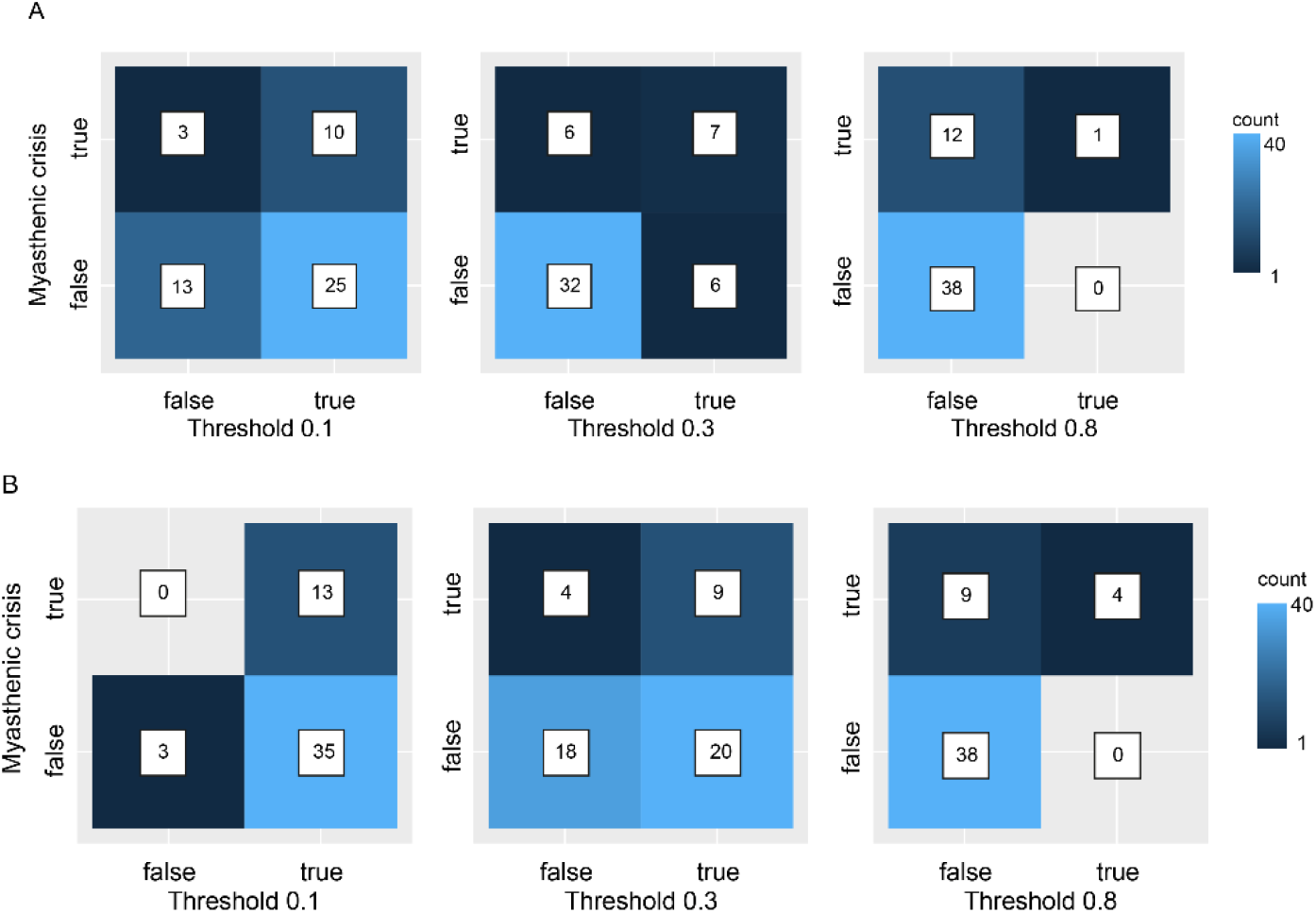
Confusion matrices of the classification results for the **(A)** Lasso regression model, and the **(B)** random forest model. *Prediction on the x-axis and ground truth on the y-axis*.

**Fig. 3:**
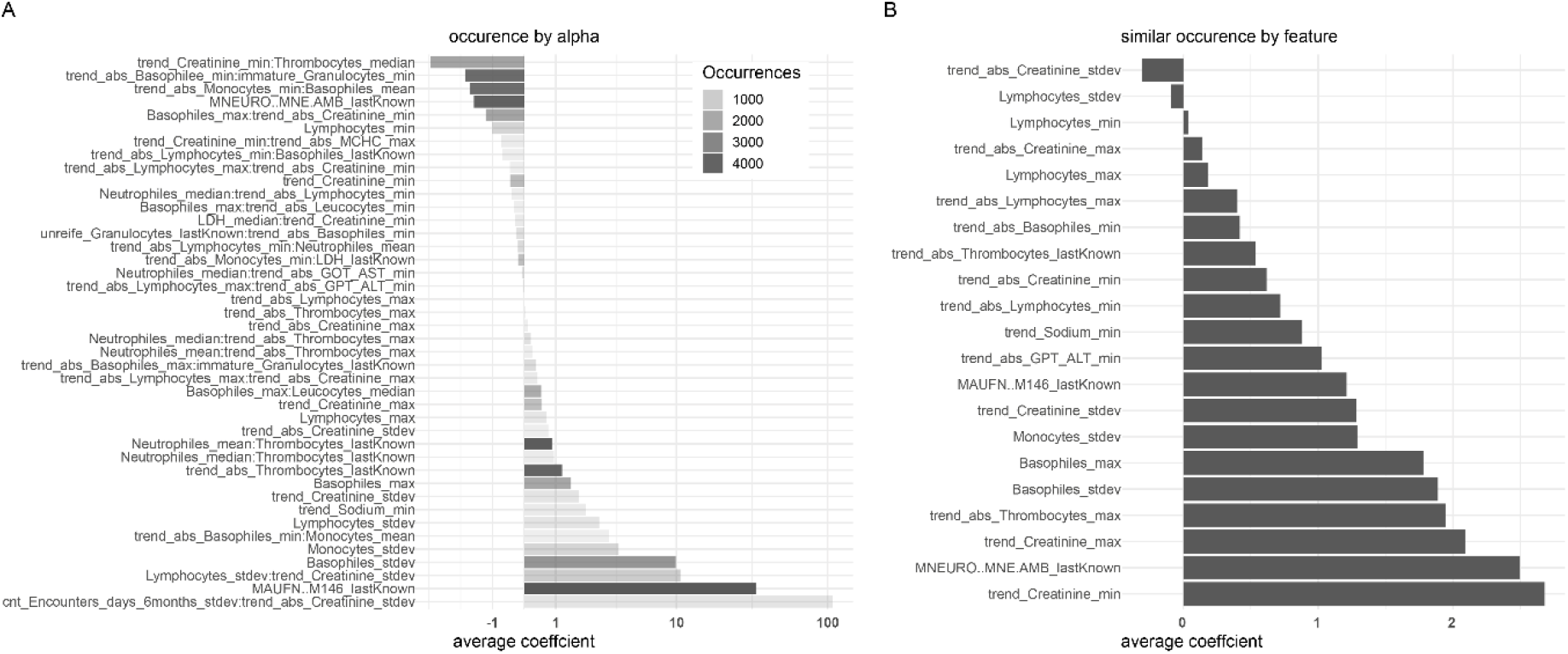
Feature importance according to mean coefficient across 100 predictions. **(A)** Feature importance for the Lasso regression. The occurrence by feature varies, because of the L1 regularization. The shading of the bars indicates the number of runs in which the feature was considered important enough (light grey = few runs, dark grey = many runs). In this context, feature importance is measured with negative or positive correlation with the dependent variable. The values around 0 are the least important ones. **(B)** Feature importance for the random forest prediction scaled to −1 to 3. In this case, the feature importance is calculated by comparing the prediction results after leaving out a feature. In this case, the negative values indicate that a feature is harmful to the prediction.

### Predictor Importance and result stability

Result stability varied by patient (Supplementary Fig. 1). However, while there were five particularly volatile prediction sets (#9, #10, #13, #20, #28) in the Lasso regression, there was only one volatile prediction set (#20) in the random forest model (Supplementary Fig. 1). This indicates that particularly the accuracy of the Lasso regression would benefit from more feature data per patient. These five patients all had an MC but were predicted as low-risk several times. We then compared the Lasso regression model with a random forest model to evaluate how much control can be asserted to the variance in the data, especially since the Lasso regression introduced variance by the chosen lambda (Supplementary Fig. 2). The high variance across runs in AUC as well as the high standard deviation of predictive score by patient across 100 predictions (Supplementary Fig. 3) suggest that using a higher number of patients as well as more data points (i.e., features) per patient would increase the prediction accuracy and reduce the number of cases when patients switch groups across runs. The chosen level of 20% imputation maximum was adequate in terms of not superseding the variance introduced by the model itself. (Supplementary Fig. 2).

Feature importance for the Lasso regression is shown with coefficients from −20 to 50 (Fig. 3A). Negative coefficients indicate anti-correlation of a feature with MC. In the Lasso regression, the features used most were the mean trend of lymphocytes (mean [sd] coefficient 191.9 [64.9]) and the path of the patient through hospital units throughout the hospital stay (mean [sd] coefficient 40.8 [14.0]) across a total of 5100 runs of the model (51 patients * 100 runs). Both were used in 99% of the runs. The most important feature in the random forest was the relative minimum trend of creatinine measurements. As the use of each feature across runs is similar in the random forest model, feature importance is measured by its accuracy coefficient after leave-one-out validation (Fig. 3B). For the random forest, the feature importance ranges from −1 to 3. Contrary to the Lasso regression, negative feature importance indicates that these features are harmful to the accuracy of the model. A full list of considered features is shown in Supplementary Table 1. Both machine learning models identified creatinine in various forms as a highly relevant feature for classification. This may suggest that multimorbidity may play a role in MC risk classification.

## Discussion

MC is the most critical presentation of MG and poses a significant burden to MG patients. It is still associated with a high morbidity, mortality, negative impact on quality of life, and requires intensive care medical treatment^4,6,7,13,14^. Thus, accurately predicting patients at risk for MC could aid treating patients preventively to avoid critical MG progression to MC as well as properly directing scarce clinical resources.

There is a growing number of studies using machine learning and artificial intelligence for research on autoimmune diseases (e.g., on type 1 diabetes^15^, multiple sclerosis^16^, rheumatoid arthritis^17^, and Crohn’s disease^18,19^), yet, in many cases focusing on genetic risk assessment. Furthermore, some applications of predictive modeling in medicine have focused on predicting Parkinson’s disease in patients before the actual clinical diagnosis^20^ or predicting the risk for exacerbation in autoimmune diseases^21^.

Although epidemiological assessment^22^, diagnosis^23^, classification of disease subtypes^24^, and therapeutic discovery^25^ for rare diseases are thought to be aided by various machine learning approaches, studies employing machine learning to monitor disease progression in MG are scarce. Further, even though disease progression models to describe disease course over time are now frequently used in drug development^26^, clinical use of disease progression modeling, e.g., to aid clinical decision making, is uncommon, particularly in rare diseases including MG. Recently, basic clinical data have been used to train a random forest classifier to predict short term clinical outcome in MG^27^. However, classifying the risk of MC based on real-world clinical data of MG patients, particularly using routine laboratory values and further case-associated data readily available at the point of care, has not been the subject of studies using machine learning for predictive modeling or risk classification.

Challenges in predictive studies in medicine are general availability of relevant clinical data, high variance in treatment procedures as well as in treatment quality across institutions and health care systems. Furthermore, in most published cases machine learning models and data have been generated specifically for the purpose of a particular prediction task in the context of a controlled trial^21,28,29^. Thus, risk prediction of new patients outside of clinical trials using real-world medical data which is generated as a part of routine treatment will be difficult.

Thus, as proof of concept, in this study we focused on using only real-world clinical data to infer patterns from patient data. We ultimately aim to make the results applicable within existing treatment procedures towards personalized disease management such as by aiding to define individual monitoring intervals or to quantify the risk of disease progression posed by a treatment change. Real-world clinical data most faithfully represents the acute disease phenotype of patients, particularly in the case of rare diseases^30^. It could also simply enhance a patient’s quality of life by easing the mental burden and adjusting the monitoring intervals^31^. Furthermore, accurate prediction of clinical exacerbation of disease using real-world clinical data aids establishing individual therapeutic concepts and tailored treatment decisions. To this end, our goal was to predict the risk for MC using two different machine learning models trained on real-world routine clinical data. Indeed, our data suggest that it is possible to discern MG patients at risk for MC from patients not at risk for MC with comparable performance as in other predictive studies on auto-immune diseases^18^. Intriguingly, the identification of creatinine, a marker of kidney injury, in various feature forms as a highly scoring features might suggest that multimorbidity, which commonly involves kidney injury, might place MG patients at high risk for MC. Indeed, multimorbidity is a known factor contributing to poor outcome in MC^4^.

Studies on risk prediction in MG usually use classical statistical models^27^ to address clinical subtypes such as thymoma patients^32–34^, specific clinical situations such as initial steroid treatment^35^ or MG subtypes classified by specific autoantibodies and prediction of factors for clinical remission. Our study used multidimensional data from a heterogenous MG cohort to learn distinctive features with the aim of predicting MC in general, and thus this work contributes a generalizable model. Furthermore, all previously available studies have a prognostic focus^31–34^, i.e., the objective is to understand which features are predictive. On the contrary, our approach is feature agnostic. We here primarily aimed at high fidelity, i.e., robust model performance maintaining reasonable group sizes as well as avoiding false negatives, in predicting whether a patient is at high risk of MC.

Classically, precision medicine has considered large scale sequencing data to tailor individual treatment decisions. Technological advances have made the use and integration of genomic, transcriptomic, and proteomic data possible^36^. While these additional data, without a doubt, retain analytic value, restricted availability of these data in a day-to-day treatment context creates a barrier between such diagnostic instruments and their utility in guiding treatment decisions. Indeed, the utility of large-scale genomic data in predicting the risk in complex sporadic conditions has been questioned^37^. Thus, advanced precision medicine should consider multimodal clinical data beyond the classical OMICS approaches. Real-world clinical data more closely reflect the medical phenotype of the patient^30^ and thereby aid understanding individual disease patterns and using individual risk factors for managing disease beyond a patient’s genotype^38^. To address this, we here investigated whether it is possible to support clinical decision making by unbiased analysis of readily available clinical data in a routine treatment setting.

Both machine learning models used herein (Lasso regression and random forest) are well understood and widely used machine learning algorithms to predict disease progression in a large variety of clinical scenarios^20,39–43^. The difference in AUC between the two models was expected, since Lasso regression was used due to the high correlation of features. The Lasso regression algorithm is known to introduce randomness (i.e., noise) because of the chosen lambda. Random forest classification was chosen specifically to control for noise in the data. In our dataset, trend features are highly predictive for the risk of MC (Fig. 3). Afterall, it is plausible to see a worsening of features in case of an imminent but not yet apparent MC. Vice versa, a patient who may be at high risk for a MC during a given visit may be in much better health (i.e., lower risk for MC) a few months later. Thus, risk for MC is not only dependent on the patient, but it can also be quite different for the same patient at two different points in time. Thus, model choice and feature design not only enabling, but focusing on a non-linear view of disease progression likely produce a more predictive result of the disease trajectory. The reduction of variance across and by patient can be addressed by including more patients as well as more data points per patient to the training set. Finally, the use of data contained in electronic health records, including laboratory parameters as features allows analyzing the reason for the good performance of the prediction algorithms. From an ethical point of view, this is a critical consideration as these models will have the capacity to alter medical decisions^44^.

### Limitations

Our findings are limited by the small sample size of 51 patients and uneven matching in some subgroups. Furthermore, for proof of concept only a selection of the available data points from the health care records were used. To prevent selection bias and better and more stable score distinction by patient, future studies will benefit from larger sample sizes as well as larger data sets including more – if not all – direct and indirect clinical parameters per patient. Furthermore, the context in which the patients live (e.g., whether patients were received nursing care or lived alone/independent or their geographic location of country vs. city) seemed to impact the outcome when reviewing the narrative of individual patient charts. Making this information machine-interpretable, would likely contribute a novel set of predictive features.

## Conclusions

This study shows that it is possible to classify the risk for MC in MG patients using longitudinal real-world clinical data. Both models showed accurate and consistent performance indicating the utility of personalized risk assessment in MG patients using machine learning models. As our models are improved, we anticipate that they will become valuable clinical tools for clinical decision support and allow unraveling the heterogeneity of MG disease phenotypes.

## Author contributions

S.B. devised and performed analytical strategies, S.B. and P.M. conceived and P.M. supervised the project, P.M. and A.M. provided clinical data, S.B. and P.M. wrote and edited the paper, A.M. edited the paper for intellectual content.

## Acknowledgments

We are grateful for support by the team of the Health Data Platform at the Berlin Institute of Health at Charité and for administrative support by S. Märschenz and S. Lischewski.

## Supplementary Data

**Supplementary Figure 1:**
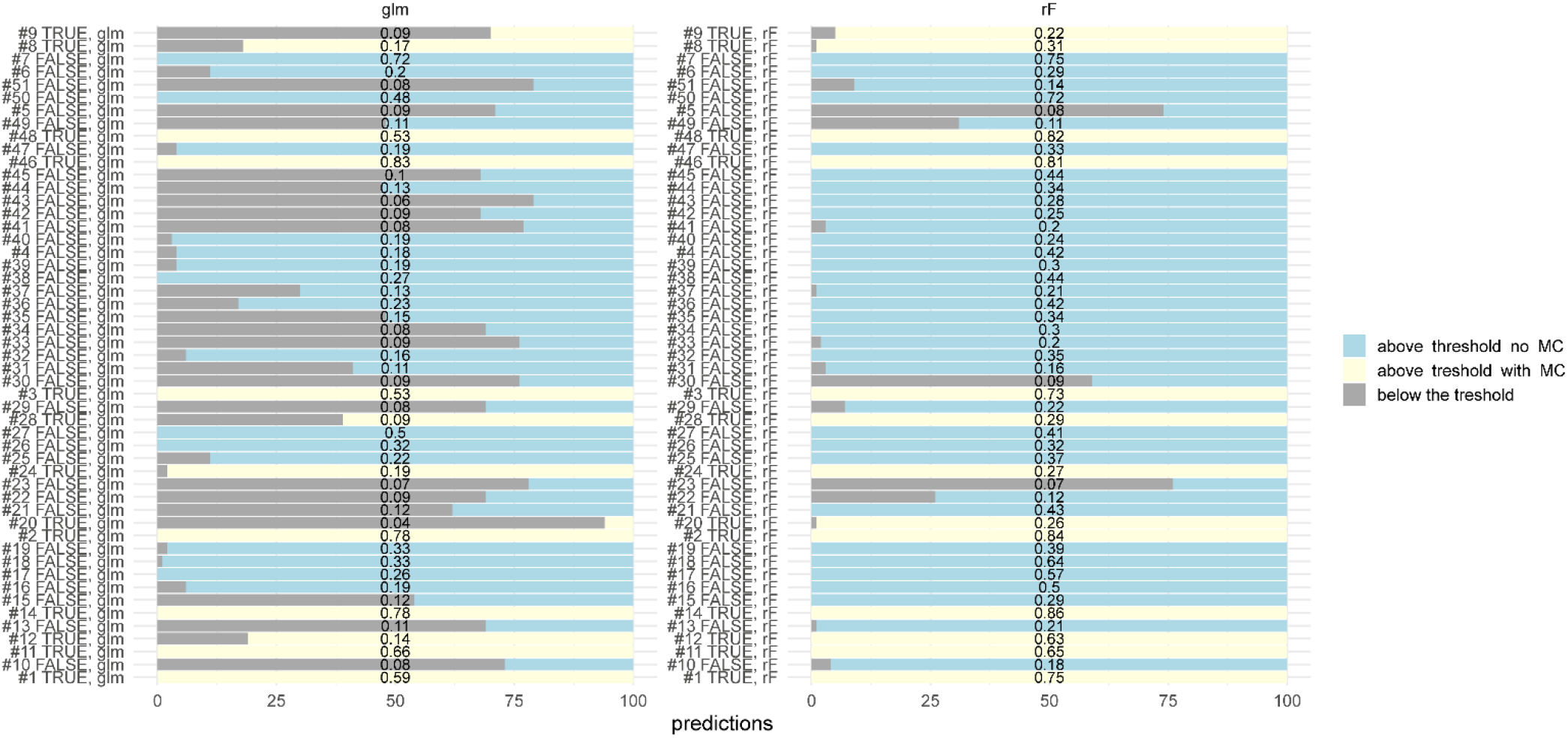
The two graphs show the stability of each patient’s predictions in the two sets of runs with different models (left: lasso regression (glm), right: random forest (rF). The black number in the middle is the mean predicted risk score for MC by patient. Each bar is a representation of all runs of the model (100 in each case). Bars that are entirely light yellow, are patients who had a crisis and were consistently predicted high risk (above the threshold (here 0.3). The bars that are light blue represent patients that were false positives, meaning they were considered high risk, but had no crisis. The high number of false positives is intended to reduce the numbers of false negatives. The dark gray bars mean that these are below the threshold and thus considered low risk. Dark gray in combination with light yellow means that these were test group members but alternated in their prediction between the groups. In both models, there are patients that were below the threshold, but had a crisis. Examples are #9, #10, #12, #13, #20, and #28 in the Lasso regression and #8, #9 and #20 in the random forest. Dark gray in combination with light blue means that these were control group members but alternated in their prediction between the groups. One interesting example for this is patient #51 in both predictions. While the patient is classified as low risk in 80% of cases in the Lasso regression model and predicted as high risk in 80% of the cases of the random forest model, this patient had no myasthenic crisis.

**Supplementary Figure 2:**
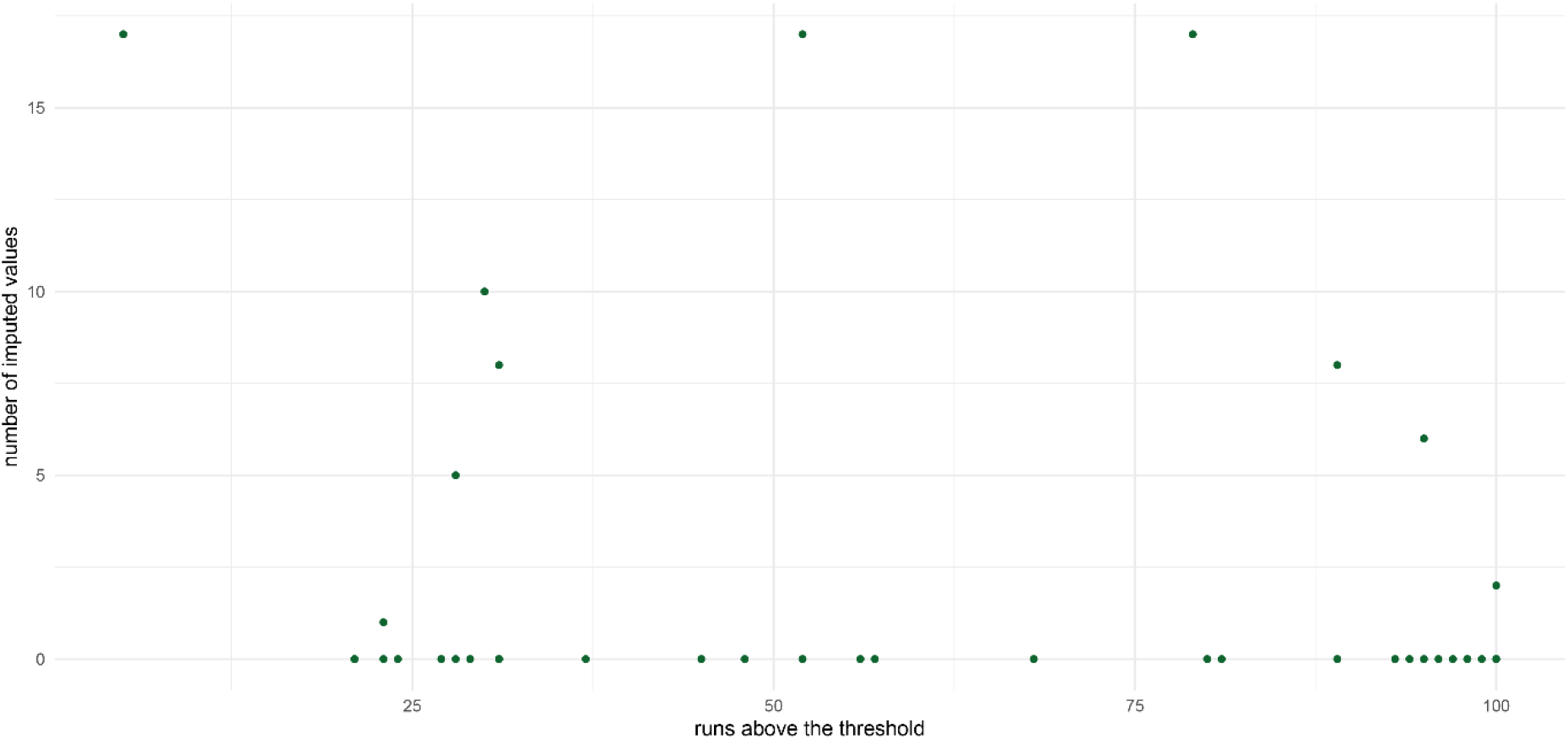
Imputed values above threshold: This graph compares the runs of the Lasso regression above the threshold with the number of imputed values by patient. One dot represents one patient. Across the number of runs we find patients with imputed values above the threshold. On the lower right of the plot there are 3 patients that were predicted above the threshold in 80-100% of the cases. At the same time, they had up to 8 imputed values. There is no obvious relationship between “predicted above the threshold” and the number of imputed values. This suggests that most of the variance by patient stems from the Lasso regression’s feature selection process. Thus, selecting a model less impacted by randomness such as the random forest is likely to show better results.

**Supplementary Figure 3:**
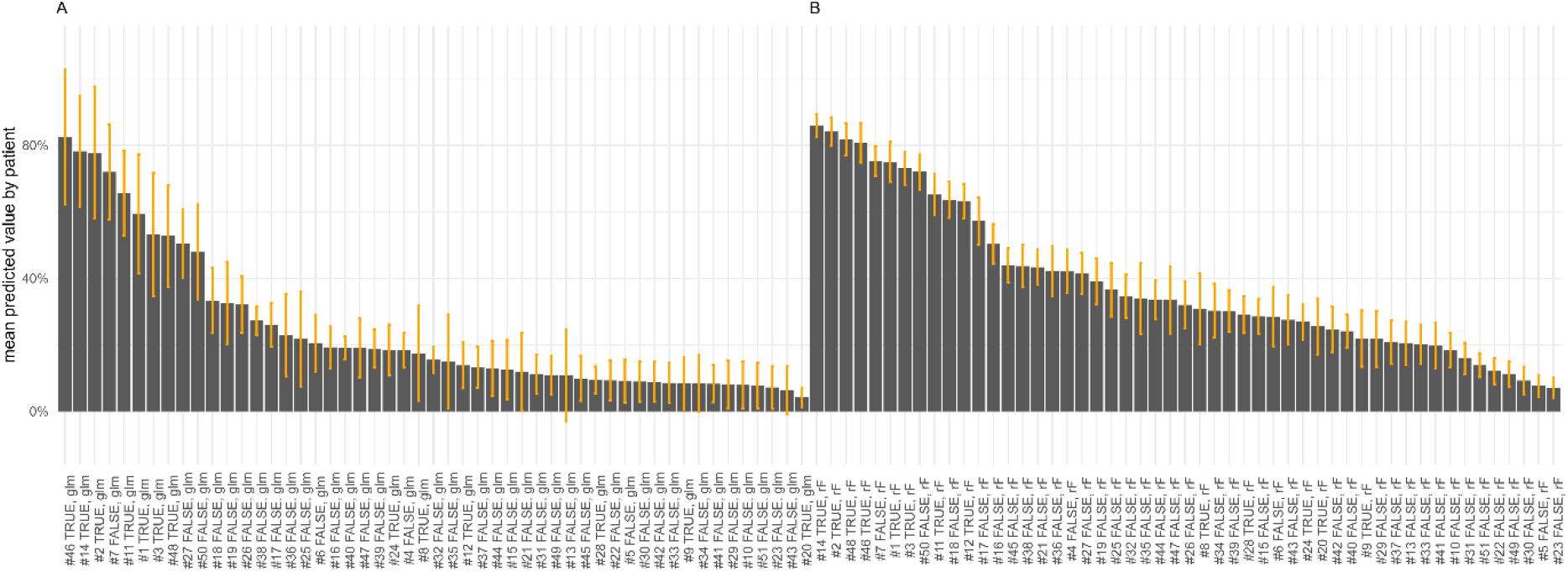
This plot shows the mean predicted value by patient in 100 runs. A) Results by patient for the Lasso regression, B) results by patient for the random forest model. The orange bar represents the standard deviation by patient. High variation suggests that both models would benefit from more in-depth data by patient. For instance, one difference that seems relevant is whether patients receive professional nursing care or live in care facilities. However, in our data set, such data are currently only available from written physicians’ notes which at present cannot quantitatively be analyzed.

**Supplementary Table 1.** List of all considered features.

| Feature Type | Feature Name |
| --- | --- |
| Patient Demographics | Age |
|  | Sex |
|  | Age of Onset |
|  | Thymus Pathology |
| Treatment Data | # Encounter Days |
|  | Hospital Path |
|  | Medication History |
| Blood Work | MCHC |
|  | GOT AST |
|  | Potassium |
|  | Sodium |
|  | Leucocytes |
|  | MCH |
|  | Creatinine |
|  | Thrombocytes |
|  | Urea |
|  | Magnesium |
|  | GPT ALT |
|  | Alkaline phosphatase |
|  | Lymphocytes |
|  | Monocytes |
|  | Basophiles |
|  | Neutrophiles |
|  | immature granulocytes |
|  | Eosinophiles |
|  | LDH |
|  | CRP |
|  | Creatinkinase CK |
|  | Titin Ab |
|  | Hct |
|  | tHB |
|  | Ca Channel PQ Type Ab |
|  | Procalcitonine |
|  | Myoglobine |
|  | Thrombo Exakt Tube |
|  | Rheumatoid factor IgM |
|  | Rheumatoid factor IgA |
|  | CK MB |
|  | GLDH |
|  | ACPA |
| **Treatment procedure** | PET/CT/MRT |
|  | Lumbar CSF puncture |
|  | EMG |
|  | Whole-body plethysmography |
|  | Thymus Excision und Resection |
|  | Ventilation |
|  | Monitoring |
|  | Complex intensive care treatment |
|  | Plasma transfusion |
|  | SSS Therapy |
|  | Immunotherapy/Immunosuppression |
|  | FPM |
|  | Tracheobronchoscopy |
|  | Blood transfusion |
|  | esophagogastroduodensoscopy |
|  | catheter |
|  | EEG |
|  | Neurography |
|  | TEE |
|  | Plasmapheresis |
|  | Sonography |
|  | Co-Diffusions capacity determination |
|  | Minimally invasive technique |
|  | Robotics/Telemedicine |
|  | Biopsy |
|  | Coloscopy |
|  | Anesthesia |
|  | FRC |
|  | requiring care |
|  | Psychosocial intervention |
|  | Chemotherapy |
|  | FSSEP/SSEP |
|  | Immunoadsorption |
|  | IM10701 |
|  | IM10671 |
|  | IM10691 |
|  | Granulocyte-stimulation |
|  | Hemodialysis/Hemofiltration |
|  | IM10711 |

**Supplementary Table 2.** R-packages used and their version.

| Package Name | Version |
| --- | --- |
| Plyr | 1.8.6 |
| Pillar | 1.5.1 |
| Tidyr | 1.1.3 |
| Stringr | 1.4.0 |
| lubridate | 1.7.10 |
| data.table | 1.14 |
| readr | 1.4.0 |
| DBI | 1.1.1 |
| dbplyr | 2.1.0 |
| RMySQL | 0.10.20 |
| purrr | 0.3.4 |
| forcats | 0.5.1 |
| readxl | 1.3.1 |
| tibble | 3.1.0 |
| mice | 3.13.0 |
| partykit | 1.2-13 |
| broom | 0.7.5 |
| rgdal | 1.5-23 |
| sp | 1.4-5 |
| scales | 1.1.1 |
| dplyr | 1.0.5 |
| plotrix | 3.8-1 |
| glmnet | 4.1-1 |
| knitr | 1.31 |
| rmarkdown | 2.7 |
| ROCR | 1.0-11 |
| glue | 1.4.2 |
| gt | 0.2.2 |
| mlr | 2.19.0 |
| pROC | 1.17.0.1 |
| libcoin | 1.0-8 |
| ggthemes | 4.2.4 |
| leaflet | 2.0.4.1 |
| ggplot2 | 3.3.3 |
| ggrepel | 0.9.1 |
| Mvtnorm | 1.1-1 |
| ParamHelpers | 1.14 |

